# Clinical Electroencephalography Findings and Considerations in Hospitalized Patients with Coronavirus SARS-CoV-2

**DOI:** 10.1101/2020.07.13.20152207

**Authors:** Neishay Ayub, Joseph Cohen, Jin Jing, Aayushee Jain, Ryan Tesh, Shibani S. Mukerji, Sahar F. Zafar, M. Brandon Westover, Eyal Y. Kimchi

## Abstract

**Background and Purpose:** Reports have suggested that severe acute respiratory syndrome coronavirus 2 (SARS-CoV-2) causes neurologic manifestations including encephalopathy and seizures. However, there has been relatively limited electrophysiology data to contextualize these specific concerns and to understand their associated clinical factors. Our objective was to identify EEG abnormalities present in patients with SARS-CoV-2, and to determine whether they reflect new or preexisting brain pathology.

**Methods:** We studied a consecutive series of hospitalized patients with SARS-CoV-2 who received an EEG, obtained using tailored safety protocols. Data from EEG reports and clinical records were analyzed to identify EEG abnormalities and possible clinical associations, including neurologic symptoms, new or preexisting brain pathology, and sedation practices.

**Results:** We identified 37 patients with SARS-CoV-2 who underwent EEG, of whom 14 had epileptiform findings (38%). Patients with epileptiform findings were more likely to have preexisting brain pathology (6/14, 43%) than patients without epileptiform findings (2/23, 9%; p=0.042). There were no clear differences in rates of acute brain pathology. One case of nonconvulsive status epilepticus was captured, but was not clearly a direct consequence of SARS-CoV-2.

Abnormalities of background rhythms were common, and patients recently sedated were more likely to lack a posterior dominant rhythm (p=0.022).

**Conclusions:** Epileptiform abnormalities were common in patients with SARS-CoV-2 referred for EEG, but particularly in the context of preexisting brain pathology and sedation. These findings suggest that neurologic manifestations during SARS-CoV-2 infection may not solely relate to the infection itself, but rather may also reflect patients’ broader, preexisting neurologic vulnerabilities.

## Introduction

As the coronavirus disease 2019 (COVID-19) pandemic has grown, there has been concern about its possible neurologic consequences^1^. COVID-19 is caused by the severe acute respiratory syndrome coronavirus 2 (SARS-CoV-2) virus, whose presentations range from asymptomatic to severe^2^. One early series of patients with severe infection described neurologic findings of impaired consciousness (14.8%) and seizure (1.1%)^3^, while another noted that 27% of patients had seizure risk factors, though clinical seizures appeared rare^4^. However, routine EEG data to support or contextualize these concerns has been limited to smaller series^5–8^, with some series abstaining from EEGs altogether due to safety concerns^4^, including limited personal protective equipment (PPE), risks of technician exposure, and possible iatrogenic spread through personnel or equipment contamination. A paucity of EEGs could underestimate the rate of seizures and epileptiform activity in SARS-CoV-2, given reports of seizures in other coronavirus outbreaks^9^ and reports of patients with SARS-CoV-2 with nonconvulsive or focal status epilepticus^10,11^. It is therefore imperative to understand what EEGs might reveal in a broader series of patients with SARS-CoV-2.

Concerns have also been raised about the neuroinvasive potential of SARS-CoV-2^12^. In some cases, the presence of neurologic symptoms has even been used to infer central nervous system (CNS) invasion^13^, and there is at least one possible case of meningoencephalitis with a positive cerebrospinal fluid (CSF) reverse-transcriptase polymerase chain reaction (RT-PCR)^14^. Autopsy series, however, have been conflicting regarding rates of neuroinvasion^15^, with the largest series suggesting that neuroinvasion is limited^16^. It is therefore unclear whether seizures or encephalopathy in SARS-CoV-2 are due to new CNS injury, reflect increased sensitivity in patients with preexisting vulnerabilities, or are associated with aspects of clinical care such as sedation.

We may be able to disambiguate this etiologic complexity by determining the burden and clinical associations of EEG abnormalities in SARS-CoV-2. We therefore undertook a study of all patients at our institution with SARS-CoV-2 who received an EEG, to test the hypothesis that there is a moderate rate of epileptiform discharges in patients with SARS-CoV-2, but that they are most common in patients with preexisting, rather than new, brain pathology.

## Methods

### Patient Cohort

We retrospectively identified all inpatients who received an EEG at an academic medical center in Boston, Massachusetts, starting after identification of the second case of COVID-19 statewide (March 2, 2020 – May 6, 2020). Patients receiving EEGs were cross-referenced against a centralized hospital database of COVID-19 status and SARS-CoV-2 results, detected using RT-PCR from nasopharyngeal samples. All patients with at least one positive SARS-CoV-2 result and a clinically interpreted EEG were included. To minimize bias, there were no further inclusion or exclusion criteria. The sample size was determined by the number of cases during the review period.

### Protocol Approvals and Patient Consents

This study of human subjects was approved by the Institutional Review Board (IRB) at Massachusetts General Hospital (Boston, MA), including review of EEG and other clinical data. The Partners Healthcare Human Research Committee provided a waiver of consent for this retrospective study.

### EEG Recordings

Protocols were developed to perform EEGs in the context of institutional guidance on care for patients with COVID-19, consistent with the American Clinical Neurophysiology Society’s recommendations^17^. EEGs were recorded using disposable Ag/AgCl scalp electrodes (Rhythymlink) using standard international 10-20 electrode placement. Care was taken with the angle of the head of the bed and manipulation of intubated patients was minimized. Although electrode placement in prone patients was anticipated, with a plan to use disposable printed electrodes and related supplies (Spes Medica TechSystem), no patient was prone at the time of electrode placement; though two were proned prior to removal. Single use bags were made for all items including skin prep (NuPrep) and conductive paste (Ten20). No air hoses or collodion were used. Leftover supplies were kept in patient rooms for possible later use and to minimize cross-contamination. Activation procedures, such as photic stimulation and hyperventilation, were not performed. Following use, computers and devices were wiped down using germicidal disposable wipes (Sani-Cloth). Portable EEG equipment was kept in a separate room for at least one hour after cleaning, away from people and other EEG machines.

All EEGs for patients with SARS-CoV-2 were performed by a single, experienced EEG technician, who sequestered at least two meters away from all other staff and had an exclusive computer. EEG technicians worked on a limited hour basis in the hospital depending on daily inpatient needs. All hospital employees were required to wear a surgical mask for their entire shift while on-site, except when working in private individual offices or in areas where employees were reliably separated by more than two meters. Additional precautions in rooms of patients with known or suspected COVID-19 included an N95 respirator, gloves, gown, and eye protection (goggles or face shield). All hospital employees had to attest daily to not having COVID-19 related symptoms, including fever, sore throat, new cough, new nasal congestion, muscle aches, new loss of smell, or shortness of breath.

### EEG Interpretation

EEG recordings were reviewed by two clinical electroencephalographers (one board-certified attending physician and one fellow) before reports were finalized in the electronic medical record. Electroencephalographers interpreted recordings remotely via secure Internet connections, which also allowed access to routine clinical data. Clinical disagreements were resolved through virtual discussion.

### Medical Record Review

Clinical EEG reports were reviewed by two neurologists to identify various possible abnormalities (listed in Table 3), including Background/Rhythm abnormalities, Rhythmic and Periodic Patterns, Sporadic Discharges, and Seizure activity. The characteristics of sporadic and periodic discharges were recorded, including whether they were identified as focal, multifocal, or generalized, and with triphasic morphology present or absent. If more than one morphology was reported then all were extracted. Patients with epileptiform findings were those with status epilepticus or seizures, with burst suppression with epileptiform activity, and with focal, multifocal, or generalized discharges, without triphasic morphology present.

**Table 1.** Clinical Characteristics of Patients with SARS-CoV-2 undergoing EEG

| **Patient Characteristics** | **(N = 37 Patients)** |
| --- | --- |
| Age in years, median (IQR) | 66.0 (59.5 - 76.3) |
| **Gender** | **n/Total (%)** |
| Female | 10/37 (27%) |
| Male | 27/37 (73%) |
| **Preferred Language** | **n/Total (%)** |
| English | 24/37 (65%) |
| Spanish | 8/37 (22%) |
| Haitian-Creole | 3/37 (8%) |
| Other | 2/37 (5%) |
| **Race/Ethnicity** | **n/Total (%)** |
| Hispanic/Latinx | 9/37 (24%) |
| Non-Hispanic Black | 12/37 (32%) |
| Non-Hispanic White | 14/37 (38%) |
| Non-Hispanic Asian | 2/37 (5%) |
| **Epidemiological Risk Factors** | **n/Total (%)** |
| Age >= 65 y | 21/37 (57%) |
| Pre-existing Lung Disease | 9/37 (24%) |
| Diabetes Mellitus | 16/37 (43%) |
| Chronic Kidney Disease | 10/37 (27%) |
| Hypertension | 23/37 (62%) |
| Cardiovascular Disease | 10/37 (27%) |
| Obesity | 14/37 (38%) |
| Immunosuppression | 5/37 (14%) |
| Living in Facility or Undomiciled | 13/37 (35%) |
| At least one risk factor above | 33/37 (89%) |
| **Respiratory Considerations** | **n/Total (%)** |
| COVID-19 Findings on Chest Imaging | 32/37 (86%) |
| Intubated | 28/37 (76%) |
| **Timing of Care** | **Median (IQR)** |
| Days from Symptoms to Admission | 5.0 (0.0 – 9.0) |
| Days from Symptoms to EEG | 17.5 (8.5 – 27.5) |
| Length of Hospital Admission (days) | 26.5 (12.5 – 41.0) |
| **Disposition** | **n/Total (%)** |
| Death or Hospice | 10/37 (27%) |
| Discharge to Facility | 23/37 (62%) |
| Discharge to Home | 3/37 (8%) |
| Continued inpatient care | 1/37 (3%) |

**Table 2.** Neurologic Context for EEG Studies in Patients with SARS-CoV-2

| **Neurologic Characteristics** | **n/Subset (%)** |
| --- | --- |
| Past Seizure History | 1/37 (3%) |
| Past History of CNS Pathology | 8/37 (22%) |
| Past Psychiatric History | 16/37 (43%) |
| Altered Mental Status in the absence of sedation | 28/37 (76%) |
| Anosmia | 4/37 (11%) |
| New Focal Exam Finding | 3/37 (8%) |
| Known Acute CNS Injury | 9/37 (24%) |
| Empiric Anti-Epileptic Drug prior to EEG | 11/37 (30%) |
| **Neuroimaging Performed** | **35/37 (95%)** |
| Computed Tomography (CT) | 35/37 (95%) |
| Magnetic Resonance Imaging (MRI) | 9/37 (24%) |
| MRIs performed with Gadolinium Contrast demonstrating Meningeal Enhancement | 0/7 (0%) |
| Neuroimaging Acutely Abnormal | 9/35 (26%) |
| **Cerebrospinal Fluid (CSF) Testing** | **4/37 (11%)** |
| CSF Studies with Abnormal WBC or Protein | 3/4 (75%) |
| CSF Studies with Abnormal WBC (>5 cells/μL) | 2/4 (50%) |
| CSF Studies with Abnormal Protein | 1/4 (25%) |
| **Reason for EEG** | **n/Subset (%)** |
| Possible Seizure | 11/37 (30%) |
| Altered Mental Status | 24/37 (65%) |
| Cardiac Arrest | 2/37 (5%) |
| **Sedation Context** | **n/Subset (%)** |
| Recent sedation | 27/37 (73%) |
| Propofol | 19/27 (70%) |
| Dexmedetomidine | 13/27 (48%) |
| Midazolam or Lorazepam | 8/27 (30%) |
| Ketamine | 2/27 (7%) |
| Sedation targeted for ventilator dyssynchrony | 13/27 (48%) |
| Sedation decreased during EEG | 10/27 (37%) |

**Table 3.**
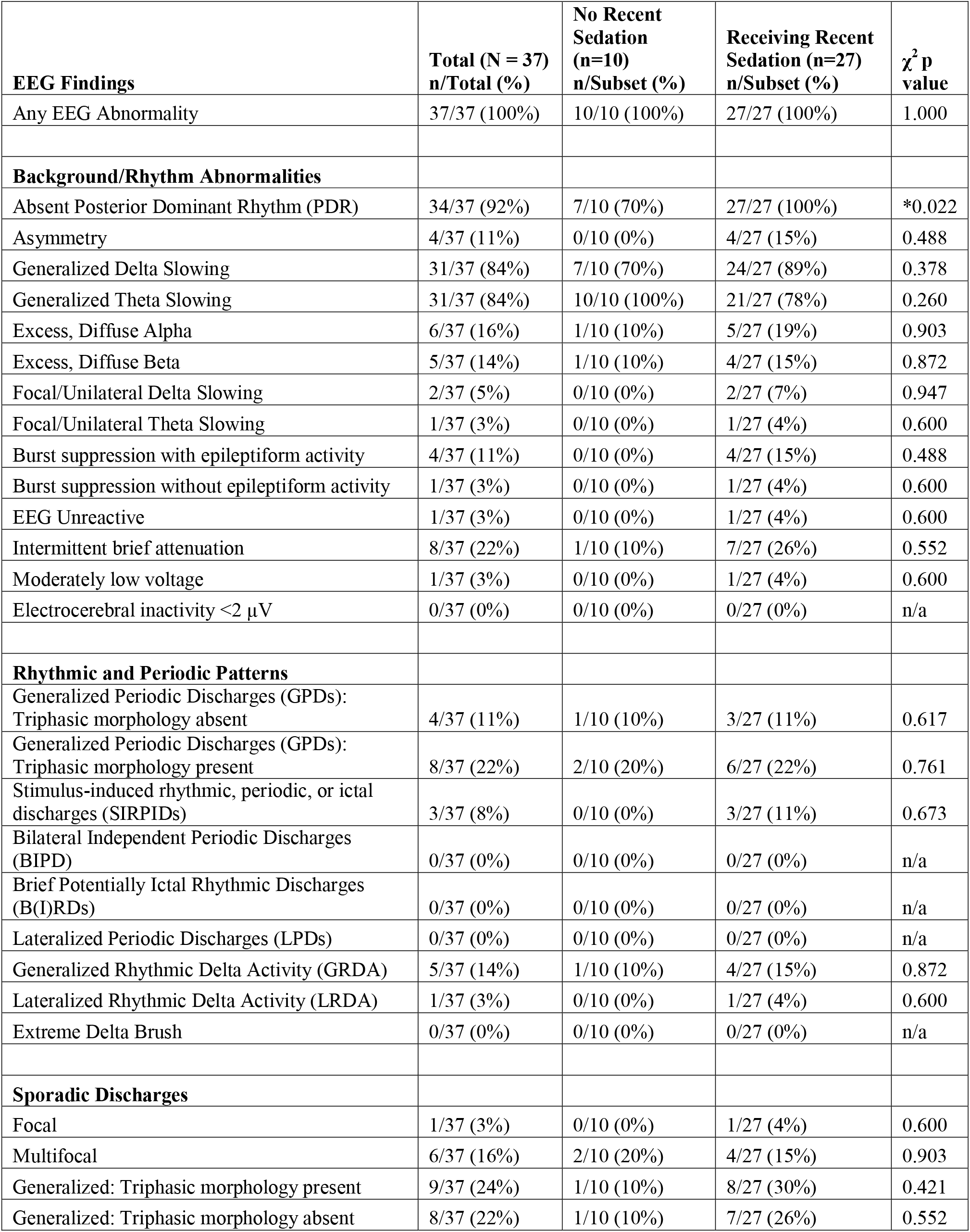
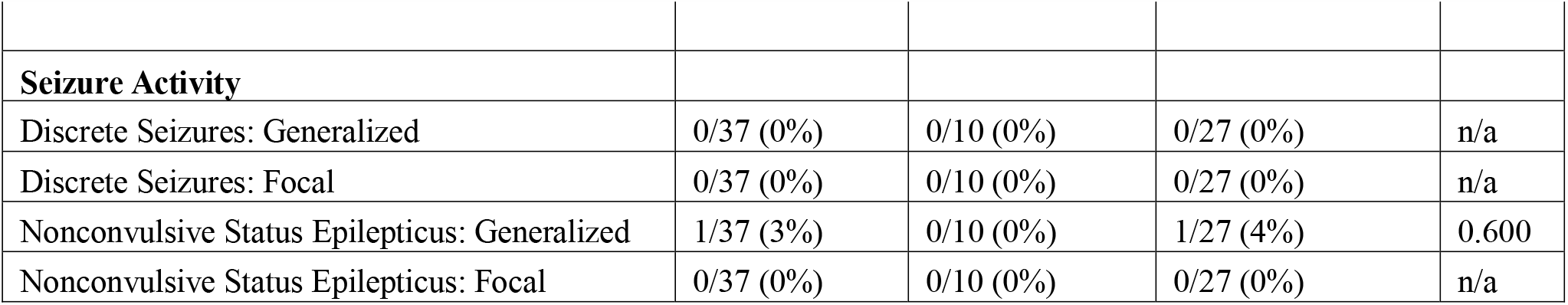
EEG Findings in Patients with SARS-CoV-2

Medical records were reviewed to extract clinical information including age, dates of symptom onset and hospitalization, medications, epidemiological risk factors^18^, bedside assessments of arousal and sedation (Richmond Agitation Sedation Scale, RASS)^19^, neurologic exams, chest and brain imaging, and initial COVID-19 related labs. We also noted the clinical questions prompting EEG. Any disagreements were resolved through virtual discussion.

### Statistical Analysis

Proportions, medians, and inter-quartile ranges (IQR) were calculated for descriptive analyses. Quantitative data was compared using rank-sum tests, and proportions were compared using Pearson Chi-square (χ^2^) tests with Yates’ continuity correction. The significance level for all tests was p<0.05. No corrections for multiple comparisons were performed given the exploratory nature of the study. Analyses were performed in MATLAB (R2018b, MathWorks).

## Results

### Patient Characteristics

We studied EEG findings in hospitalized patients with SARS-CoV-2. A total of 307 patients received an acute care EEG during the study period, of which 37 tested positive for SARS-CoV-2, all before EEG. From the date of the first EEG in a patient positive for SARS-CoV-2 (3/27/2020), 25% of subsequent EEGs were in patients with SARS-CoV-2 (37/149 patients). The patients’ clinical characteristics are displayed in Table 1. Most patients had chest imaging findings consistent with COVID-19 (32/37, 86%), and most were intubated (28/37, 76%). Laboratory studies revealed elevated inflammatory markers (Supplemental Table 1).

### Neurologic Context for EEG Studies

Patients had diverse neurologic characteristics (Table 2). Only one patient had a history of epilepsy prior to presentation (1/37, 3%), secondary to prior intraparenchymal hemorrhage. Seven other patients had a history of central nervous system (CNS) disease. One patient had Dementia with Lewy Bodies, and six patients previously had ischemic strokes, including one in the context of cerebral aneurysm coiling and one in the context of metastatic small cell lung cancer. Acutely, most patients had altered mental status, even in the absence of sedation or persistently during sedation weans (28/37, 76%). Some patients were reported to have anosmia (4/37, 11%). New focal signs were relatively rare: two had patients had gaze preferences and one had hemiparesis (3/37, 8%).

Based on clinical evaluation, neuroimaging, and cerebrospinal fluid (CSF) studies, nine patients were determined to have acute CNS pathology (9/37, 24%), including three with ischemic stroke, three with intracerebral hemorrhage, and one each with cryptococcal meningitis in the setting of virally unsuppressed HIV infection, paraneoplastic encephalitis in the setting of small cell lung cancer, and subacute combined degeneration in the setting of nitrous oxide abuse. The latter three non-vascular events were thought not to be direct results of SARS-CoV-2 per care teams.

Several concerns prompted EEG, including convulsions with unresponsiveness concerning for seizures (11/37, 30%), altered mental status (24/37, 65%), and monitoring during post-cardiac arrest hypothermia (2/37, 5%). Most EEGs involved long-term monitoring (LTM; 23/37, 65%), with a median of 22.5 hours (IQR 0.4 – 43.4 hours). Most patients had received recent sedation, either on the day of EEG or the day prior (27/37, 73%), and patients had a median RASS of −3 (IQR −4 to −2). Only 10 patients could have sedation weaned during EEG. Sedation was often specifically targeted to deep levels due to ventilator dyssynchrony in the context of acute respiratory distress syndrome (ARDS) (13/27 deeply sedated for dyssynchrony, 48%).

### EEG Results

EEG revealed findings in all patients (Table 3). The most common EEG findings were generalized abnormalities of background rhythms, including absent posterior dominant rhythms (34/37, 92%) and generalized delta and theta slowing (Figure 1A). Due to the possible effects of sedation on background rhythms, we stratified patients according to whether they had recently received sedation. Patients who had recently received sedation were more likely to lack a posterior dominant rhythm (absent in 27/27, 100%), compared to 7/10 (70%), of patients who had not recently received sedation (χ^2^ p=0.022).

**Figure 1.**
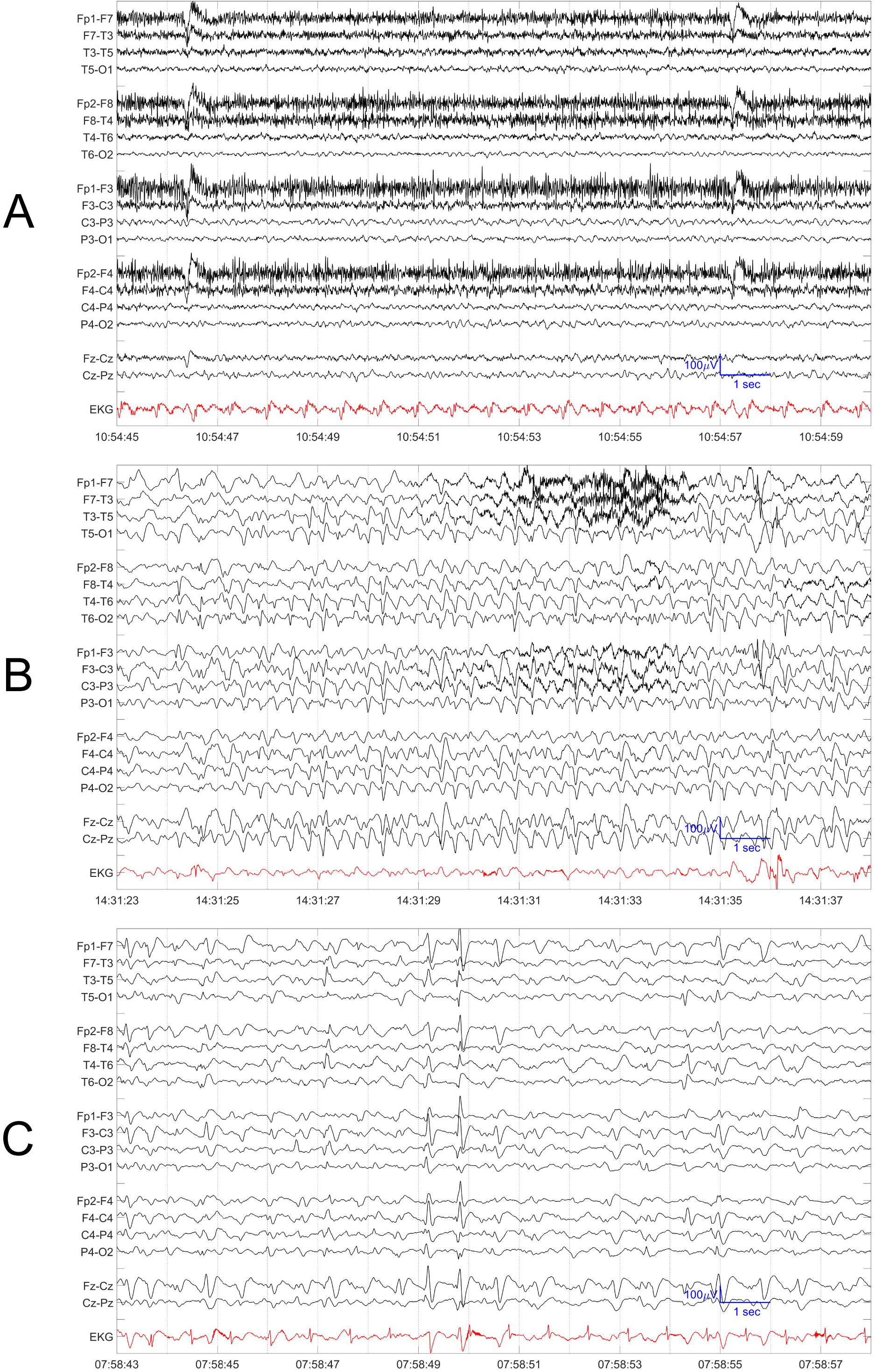
Representative EEGs from Patients with SARS-CoV-2 (A) Example EEG of a patient with SARS-CoV-2 who was not intubated and not receiving generalized sedation, with absent posterior dominant rhythm and the presence of generalized theta slowing. For all panels, data was filtered with a bandpass filter between 1-70Hz and a notch filter at 60 Hz. EEG is shown in black in a longitudinal bipolar montage, EKG is shown in red. The sensitivity is indicated in blue for each panel. (B) Example EEG of a patient with SARS-CoV-2 who had nonconvulsive status epilepticus as described in the Results, with generalized periodic discharges with sharp wave morphology at a frequency of 3-4 Hz when propofol was held. (C) Example EEG of a patient with SARS-CoV-2 with multiple discharge morphologies, including both triphasic and otherwise generalized sharp wave morphologies.

EEG studies in seven patients included capture of convulsive or concerning events, but none had electrographic correlates of discrete seizures. Electrographic seizures were also uncommon, with only one case of nonconvulsive status epilepticus (2.7%; 95% confidence interval 0-7.9%). This patient was a woman in her 60s with a history twenty years prior of a left posterior communicating artery aneurysm, whose coiling was complicated by a left precentral gyrus infarct. She had no prior history of seizures. She presented acutely with syncope, and Chest CT revealed an ascending aortic dissection with rupture and tamponade, but no signs of viral pneumonia. She tested positive for SARS-CoV-2. She underwent emergent repair, and aortic pathology revealed focal chronic inflammation composed primarily of CD3+CD4+ T-cells and CD68+CD163+ macrophages, but no diagnostic features of active aortitis. Post-operatively she developed left sided head and arm twitching while being weaned from propofol. CT did not reveal acute pathology. Her EEG demonstrated right frontal and generalized periodic sharp waves, without convulsive activity but with frequency increasing to 4 Hz when propofol was held, which was determined to be electrographically and clinically consistent with non-convulsive status epilepticus (Figure 1B). After treatment of non-convulsive status epilepticus, an MRI revealed small acute infarcts in right middle frontal gyrus and left superior parietal lobule, consistent with emboli from her dissection. Following care, she was discharged to a rehabilitation facility.

Sporadic and periodic discharges were also observed in the EEG studies (Figure 1C). Discharges were morphologically classified by clinical EEG readers, and patients sometimes had more than one morphology. There was no definite frontal or temporal predominance. We identified patients with epileptiform findings as those with focal, multifocal, or generalized discharges, excluding those with triphasic morphology, as well as patients with electrographic seizures, status epilepticus, or burst suppression with epileptiform activity. 38% of patients had epileptiform findings (14/37, Table 4). We evaluated what clinical characteristics were more common in patients with epileptiform findings. A history of CNS pathology was more common in patients with epileptiform findings (6/14 patients with epileptiform findings had a history of CNS pathology, compared to 2/23 patients without epileptiform findings, χ^2^ p=0.042). EEGs with epileptiform findings were also obtained earlier in patient’s courses (p=0.044).

**Table 4.** Clinical Associations with Epileptiform Findings in Patients with SARS-CoV-2

|  | EEGs without Epileptiform Findings (N=23) | EEG with Epileptiform Findings (N=14) |  |
| --- | --- | --- | --- |
| Patient Characteristics | Median (IQR) | Median (IQR) | Rank-sum p value |
| Age (years) | 66.0 (57.0 - 78.0) | 66.5 (63.0 - 73.0) | 0.888 |
| Days from Symptoms to EEG | 20.0 (10.8 – 28.0) | 4.0 (1.0 – 16.5) | *0.044 |
| Patient Characteristics | n/Subset (%) | n/Subset (%) | $\chi^2$ p value |
| COVID-19 Findings on Chest Imaging | 21/23 (91%) | 11/14 (79%) | 0.547 |
| Past History of CNS Pathology | 2/23 (9%) | 6/14 (43%) | *0.042 |
| Anosmia | 4/23 (17%) | 0/14 (0%) | 0.269 |
| Altered Mental Status in the absence of sedation | 17/23 (74%) | 11/14 (79%) | 0.940 |
| Known Acute CNS Injury | 6/23 (26%) | 3/14 (21%) | 0.940 |
| Recent Sedation | 14/23 (61%) | 7/14 (50%) | 0.760 |
| Reason for EEG | n/Subset (%) | n/Subset (%) | $\chi^2$ p value |
| Possible Seizure | 5/23 (22%) | 6/14 (43%) | 0.321 |
| Altered Mental Status | 17/23 (74%) | 7/14 (50%) | 0.262 |
| Cardiac Arrest | 1/23 (4%) | 1/14 (7%) | 0.700 |
| Impact of EEG | n/Subset (%) | n/Subset (%) | $\chi^2$ p value |
| Antiepileptic Drug Started | 0/23 (0%) | 2/14 (14%) | 0.265 |
| Benzodiazepine Trial Initiated | 0/23 (0%) | 2/14 (14%) | 0.265 |

Care was escalated in four patients with epileptiform findings. Two patients received increased antiepileptic therapy. Two additional patients received benzodiazepine trials for generalized periodic discharges, one of which improved clinically but was ultimately thought to be more consistent with catatonia than nonconvulsive status epilepticus.

### EEG Safety

To date, none of our EEG staff has tested positive for SARS-CoV-2, including the technician performing the studies, other EEG technicians, and interpreting neurophysiologists. Additionally, there have been no known cases nor safety reports in our institution regarding cross-contamination of other patients as a result of EEG testing.

## Discussion

### SARS-CoV-2 and Epileptiform EEG Findings

All patients with SARS-CoV-2 in our study who received EEGs for clinical purposes had EEG abnormalities, including epileptiform findings in just over a third of the patients, despite only one patient having a history of epilepsy. These findings have been seen in other types of critical illness^20,21^, suggesting that direct neuroinvasion need not occur to explain such findings. Epileptiform activity appeared to be associated with preexisting brain pathology, but not acute clinical or radiographic findings. Case reports of status epilepticus in SARS-CoV-2 have been described both with^11,22^ and without^10,23^ prior CNS pathology. Although several patients had acute CNS pathology, a direct effect of SARS-CoV-2 was sometimes hard to ascertain, even for our patient with nonconvulsive status epilepticus.

Epileptiform abnormalities were relatively diverse including multifocal and generalized discharges, and other potentially epileptiform findings such as discharges with triphasic morphology. While there has been concern that intranasal infection may be a potential route for coronavirus neuroinvasion^24^, there was no clear association between epileptiform findings and anosmia, nor was there a fronto-temporal predominance to suggest specifically limbic involvement. Other groups have noted frontally predominant abnormalities in patients with SARS-CoV-2^7,8,25,26^, but in at least one paper frontal slowing appeared similar to that seen in metabolic encephalopathies and correlated with systemic lab findings^7^. Although a few encephalitides are associated with suggestive EEG findings, such as very low frequency periodic complexes in measles-related subacute sclerosing panencephalitisis^27^ or lateralized periodic epileptiform discharges in HSV encephalitis^28^, neither finding is necessarily specific. Our study did not identify an EEG finding more specific to SARS-CoV-2 than other syndromes.

Several EEGs included capture of convulsive or concerning events, but none had electrographic correlates. This suggests the need for care regarding the diagnosis of seizures solely based on convulsions in patients with SARS-CoV-2, which can also manifest with myoclonus^29^. More detail may be necessary when seizures are diagnosed or reported in COVID-19 without description of ancillary data.

### SARS-CoV-2 and EEG Background Rhythms

In addition to epileptiform abnormalities, the most common EEG findings were background rhythms abnormalities, a nonspecific finding that can be associated with poor prognosis^30^. Some abnormalities, for example generalized theta slowing, were frequent even in patients who had not recently received sedation, and this can be seen in non-encephalitic conditions such as toxic-metabolic encephalopathies^31^. However, the increased lack of a posterior dominant rhythm in patients recently receiving sedation suggests that some background rhythm abnormalities in SARS-CoV-2 may not simply reflect direct effects of SARS-CoV-2 infection, but may include contributions from its management, such as sedation.

A consideration that was unexpected at the outset of this study was how frequently patients were deeply sedated, beyond typical sedation goals (for example to a RASS of −3, rather than −1 to 0). While ICU sedation can unintentionally become deeper than intended, especially when the number of bedside evaluations are limited^32^, sedation was also intentionally deep and rarely lifted successfully due to ventilator dyssynchrony. The risks and benefits of deep sedation in Acute Respiratory Distress Syndrome (ARDS) are complex^33^, and the sedation strategy that most effectively balances patient-specific risks and benefits in SARS-CoV-2 remains unknown. Heavy sedation is likely to affect EEG sensitivity and specificity and has not been considered in all prior SARS-CoV-2 EEG reports. Given that several of our EEGs were prompted by convulsions in the setting of weaning sedation, we recommend anticipating whether and when attempts will be made to decrease sedation during long-term EEGs.

### EEG and SARS-CoV-2 Safety Considerations

As the COVID-19 pandemic caused by SARS-CoV-2 continues, there will be increasing need to safely perform diagnostic tests that were previously routine^34,35^. To date there has been no clear transmission within or from our division. However, there is limited data to guide safety protocols and limit iatrogenic spread, including among healthcare workers^36^. Protocols are likely to depend on local PPE availability and evolving understandings of COVID-19 spread. The benefits of masking patients remains uncertain, and exhalation airflow is redirected by masks, potentially flowing superiorly, with unclear consequences for EEG technicians who often stand above patients’ heads^37^. Tracking healthcare related cases of SARS-CoV-2 would be helpful to understand risks of transmission to and from specific provider roles.

## Limitations

Our study has several imitations. This case series was drawn from a single center in order to evaluate data efficiently, albeit from a hospital with the largest caseload in a state considered one of the United States’ early epicenters. The series was also relatively small, though to our knowledge, this is the largest known series to date of patients receiving standard EEGs for any coronavirus. All EEGs were ordered at the discretion of providers, which may introduce clinical biases that may limit generalizability to patients who do not prompt such studies. Our rate of electrographic seizures and estimated confidence interval, however, agrees with other, smaller series, suggesting a ≤8% rate of seizures even in such selected populations^3,38–40^. This series included a heterogeneous population, including some patients who appeared asymptomatic from a respiratory perspective. Asymptomatic patients will likely form an increasing portion of SARS-CoV-2 studies as both viral spread and testing increase. Additionally, while we have shared the specific safety protocols undertaken by our division at this time, we anticipate that protocols will evolve as more is learned.

## Conclusion

EEG provides a dynamic window into the dramatic ways in which the minds and brains of our patients are affected by the systemic, neurologic, and psychosocial consequences of SARS-CoV-Epileptiform abnormalities were common in patients with SARS-CoV-2 referred for EEG and were associated with preexisting brain pathology. These findings suggest that the neurologic consequences of SARS-CoV-2, including seizures and encephalopathy, may not relate solely to infection itself, but also to how neurologically vulnerable patients are affected more broadly, including the ways in which we care for them.

## Data Availability

Anonymized participant data used and analyzed during the current study will be shared with reasonable request from qualified researchers.

## Acknowledgements

The authors wish to acknowledge and express their appreciation to the patients who were a part of this study, as well as the EEG technicians and clinical neurophysiologists of the MGH Division of Clinical Neurophysiology who participated in their clinical care.

## Conflict of Interest Notification

Dr. Ayub has nothing to disclose.

Mr. Cohen has nothing to disclose.

Dr. Jing has nothing to disclose.

Ms. Jain has nothing to disclose.

Mr. Tesh has nothing to disclose.

Dr. Mukerji reports grants from NIH/NIMH, grants from Harvard University, during the conduct of the study.

Dr. Zafar reports grants from NIH/NINDS, during the conduct of the study.

Dr. Westover reports grants from NIH/NINDS, from Massachusetts General Hospital, during the conduct of the study.

Dr. Kimchi reports grants from NIH/NIMH, during the conduct of the study.

## Supplementary Material

Supplemental material for this article is available online.

